# Toward Early Diagnosis and Therapeutic Discovery in CLN3 Disease: A Computational Biomarker Discovery Framework

**DOI:** 10.64898/2026.05.01.26352147

**Authors:** Shixue Sun, An N. Dang Do, Audrey Thurm, Ariane Soldatos, Qian Zhu

## Abstract

**Background:** CLN3 disease, also known as juvenile neuronal ceroid lipofuscinosis, is a rare and neurodegenerative disorder characterized by the accumulation of lipopigments in the cells, progressive cognitive decline, seizures, and vision loss. Biomarker discovery in CLN3 disease is essential for enabling early and accurate diagnosis, which is critical given its neurodegenerative course. Biomarkers provide objective measures to track disease progression, stratify patients, and serve as surrogate endpoints in clinical trials, thereby accelerating therapeutic development. They also offer valuable insights into underlying disease mechanisms and treatment response, ultimately advancing individualized medicine and improving clinical outcomes.

**Methods:** We developed various machine learning models to predict potential protein biomarkers in CLN3 disease using proteomics data and laboratory tests collected from participants in a prospective, observational cohort. To prioritize and evaluate these candidates, we conducted protein-protein interaction (PPI) network analysis and pathway enrichment, ranking proteins based on their topological importance. The top 20 proteins were selected as candidate biomarkers and corroborated using a publicly available CLN3 transcriptomic dataset. Receiver operating characteristic (ROC) curve analysis was performed to assess the discriminative power of each candidate, with AUROC values calculated to quantify their classification performance.

**Results:** Our computational approach identified six promising biomarker candidates: OSM, IL6R, LMNB1, HIF1A, NPM1, and CSF1. Among them, OSM and HIF1A showed marked differential expression in CLN3 patients, particularly those with slow disease progression. LMNB1 expression was elevated in patients with faster disease progression, suggesting its utility as a prognostic biomarker. These findings highlight the robustness of our biomarker selection, indicating that these six genes may serve as effective diagnostic markers for CLN3 disease.

**Conclusions:** Our findings demonstrate the utility of data-driven approaches for biomarker discovery in CLN3 and offer new insights into the molecular mechanisms of the disease, with broader implications for improving diagnosis and prognosis in other rare diseases.

## 1. Introduction

CLN3 disease, also known as juvenile neuronal ceroid lipofuscinosis (GARD:0005897), is a rare neurodegenerative disorder characterized by the accumulation of lipopigments in the cells, leading to cognitive decline, seizures, and vision loss in affected individuals [1]. This devastating condition primarily affects children and young adults, with initial symptom typically being declining visual ability starting between the ages of 4 and 6 years. The pathogenesis of CLN3 involves mutations in the *CLN3* gene, which encodes a protein critical for normal cellular function, and plays an emerging role in regulating levels of phospholipid-related compounds [2–6]. The prevalence of CLN3 is estimated to as 2-5 in 100,000, making it as a rare disease with limited public awareness and research attention [1, 7]. Current treatments remain symptomatic and supportive, focusing on seizure management and physical therapy, with no cure or disease-modifying therapy available [8, 9]. Several candidate therapies are currently in preclinical or early-phase clinical studies [10–14], underscoring the urgent need for identifying robust, disease relevant, and quantitative fluid biomarkers.

Biomarkers are critical for elucidating disease mechanisms, monitoring disease progression, and evaluating therapeutic responses. The presentation of signs and symptoms of CLN3 disease emerge gradually over years and the disease progression and severity are varying. Biomarkers that can inform prediction of CLN3 severity and disease progression would greatly enhance clinical care and facilitate precision treatments. However, to date, only a few studies have studied fluid biomarkers for this purpose [3, 15–19]. For example, elevated levels of glycerophosphoinositol (GPI) have been identified in both cerebrospinal fluid (CSF) and blood samples from CLN3 patients, CLN3 Δex7-8 pigs, and CLN3 Δex7-8 mice, emphasizing its potential as a highly sensitive and specific biomarker for CLN3 disease given its role in the clearance of glycerophosphodiesters from lysosomes [3, 19]. Additionally, protein biomarkers such as NEFL and CHIT1 have been found to correlate with the neurological status in CLN3, based on proteomics data from cerebrospinal fluid (CSF) samples analyzed by Proximity Extension Assay (PEA; Olink®) technology (9). Other proteins, including CHIT1, NELL1, and ISLR2, have also been identified as biomarker candidates from two proteomics datasets generated by PEA and mass spectrometry (MS) (6).

Artificial Intelligence (AI) models have emerged as powerful tools in biomarker discovery, enabling extraction of disease-relevant features from omics and clinical data [20, 21]. By learning complex patterns and prioritizing stable predictive variables, these models support the discovery of robust candidate biomarkers for diagnosis, prognosis, and therapeutic targeting. For example, a pharmaco-metabolomics study with plasma samples applied Random Forest (RF) and Support Vector Machine (SVM) to identify predictive markers strongly associated with disease progression and treatment response for Amyotrophic Lateral Sclerosis (ALS) [22]. van der Burgh *et al*. demonstrated that combining clinical characteristics with MRI data using <u>artificial neural network</u> (ANN) algorithm significantly improved the accuracy of predicting survival prediction in ALS patients [23]. AI has also shown promise in advancing treatment strategies for rare diseases. For instance, an IBM Watson’s application in neurodegenerative disease research has identified novel RNA-binding proteins (RBPs) linked to ALS by minging literatures to establish semantic similarity with previously known ALS associated RBPs [24]. Another study applied multi-output regression machine learning methodologies to investigate the Fanconi Anemia pathway, predicting over 20 potential therapeutic targets by modeling the relationships between external proteins and disease-related cellular functions [25]. Despite these advantages, AI-driven biomarker discovery in rare diseases faces several obstacles, including limited sample sizes, high disease heterogeneity and substantial missing data [26, 27].

In this study, we introduced a computational framework to programmatically identify fluid biomarker candidates for CLN3 disease using proteomics data and clinical data derived from a prospective CLN3 observational natural history trial (NCT03307304). This framework is composed of different computational components including data processing, predictive model for biomarker prediction, and biomarker prioritization and evaluation. The results highlight the utility of AI-driven approaches in biomarker discovery in CLN3 disease and offer new insights into its underlying molecular mechanisms, with potential implications for advancing diagnosis and prognosis in other rare diseases.

## 2. Materials and Methods

In this study, we developed multiple machine learning models using proteomics data from the CLN3 clinical protocol (NCT03307304) for biomarker discovery for CLN3 disease. Figure 1 illustrates the overall study workflow.

**Figure 1.**
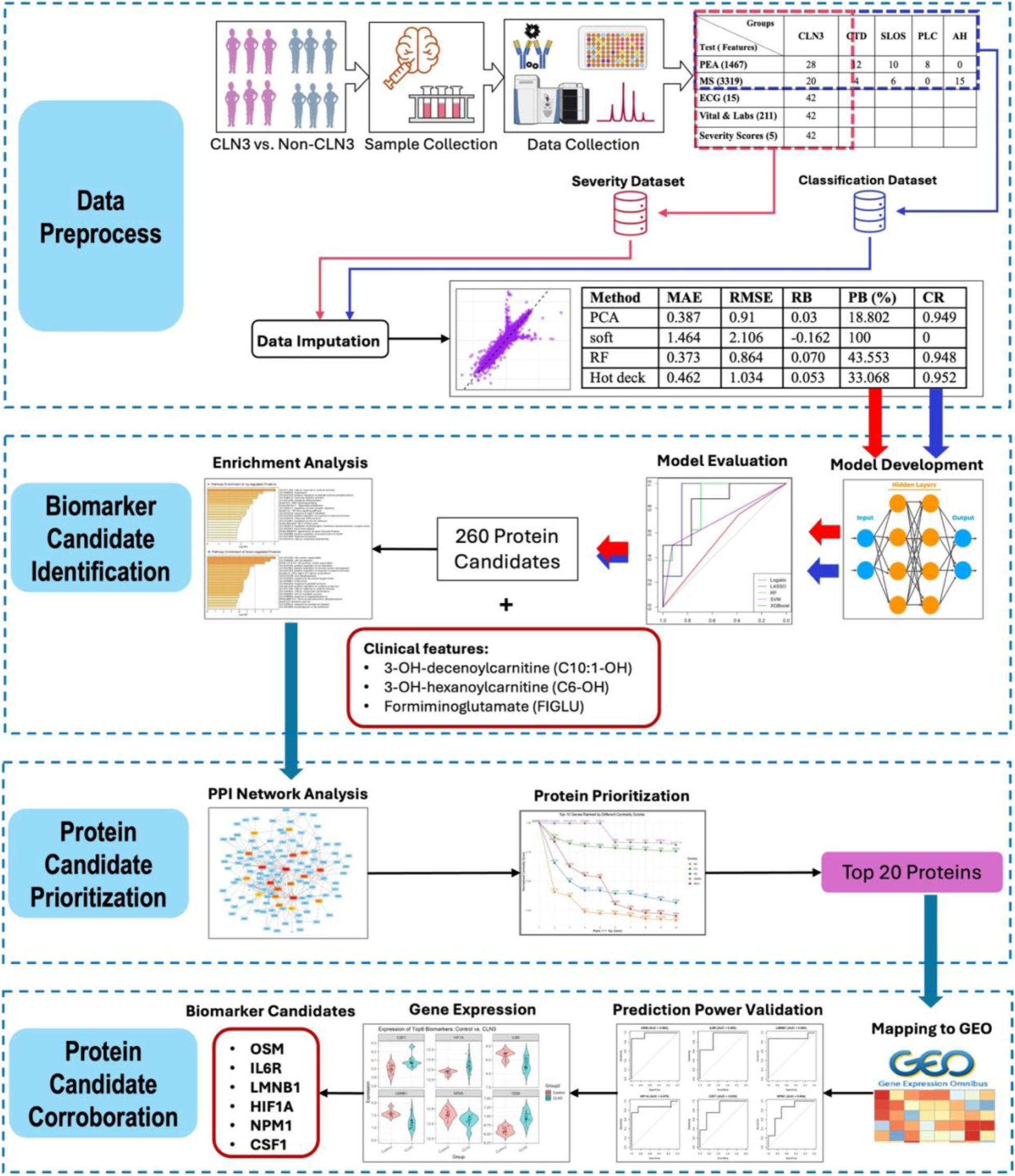
The overall study workflow of this project. (Features included in the two red boxes are the final biomarkers identified from this study including three metabolites and six protein candidates)

### 2.1 Data Preparation

We obtained proteomics data and clinical data from samples collected through NIH IRB-approved prospective, observational studies [CLN3 disease: NCT03307304. Creatine Transporter Deficiency (CTD): NCT02931682. Smith-Lemli-Opitz syndrome (SLOS): NCT00001721. NPC: NCT00344331]. Participants, parents, or guardians provided assent or consent as appropriate. Details regarding participant demographics, clinical and proteomic data can be found in our previous publications [15, 16, 18].

This study analyzed cross-sectional data collected from 42 participants with CLN3 disease enrolled from November 27, 2017 and September 25, 2023. Cerebrospinal fluid (CSF) samples were analyzed using two proteomic platforms: Proximal Extension Assay (PEA; Olink®) [28] and mass spectrometry (MS). The CSF samples from non-CLN3 participants included individuals with CTD (n=12), SLOS (n=10), residual pediatric laboratory controls (PLC, n=8), and healthy adult volunteers obtained commercially (PrecisionMed, https://www.precisionmed.com/) (AH, n=15). A total of 28 CLN3 and 30 non-CLN3 samples were assayed using PEA [28], while 20 CLN3 and 25 non-CLN3 were analyzed by MS. Notably, 20 CLN3 and 10 non-CLN3 samples were assessed using both platforms. At the time of CSF collection, CLN3 participants underwent comprehensive clinical evaluations, including vital sign, electrocardiogram, and laboratory tests (The raw data of all participants, including demographics, proteomics, ECG, laboratory tests, and clinical evaluations can be found in the Supplementary_ File_1). An overview of the study data is provided in Table 1.

**Table 1.**
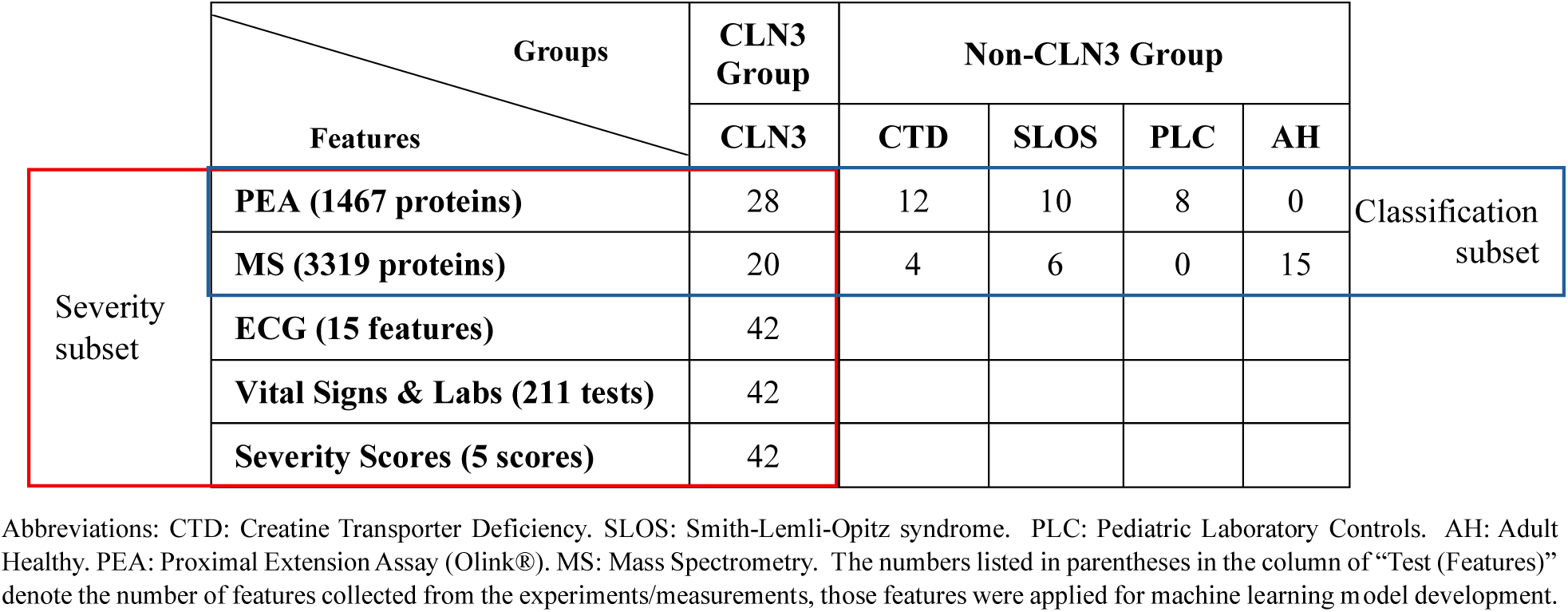
Overview of the study participants and the related datasets.

|  | Features | CLN3 Group | Non-CLN3 Group |  |  |  |  |
| --- | --- | --- | --- | --- | --- | --- | --- |
|  |  | CLN3 | CTD | SLOS | PLC | AH |  |
| Severity subset | PEA (1467 proteins) | 28 | 12 | 10 | 8 | 0 | Classification subset |
|  | MS (3319 proteins) | 20 | 4 | 6 | 0 | 15 |  |
|  | ECG (15 features) | 42 |  |  |  |  |  |
|  | Vital Signs & Labs (211 tests) | 42 |  |  |  |  |  |
|  | Severity Scores (5 scores) | 42 |  |  |  |  |  |

Abbreviations: CTD: Creatine Transporter Deficiency. SLOS: Smith-Lemli-Opitz syndrome. PLC: Pediatric Laboratory Controls. AH: Adult Healthy. PEA: Proximal Extension Assay (Olink®). MS: Mass Spectrometry. The numbers listed in parentheses in the column of “Test (Features)” denote the number of features collected from the experiments/measurements, those features were applied for machine learning model development.

Disease state was evaluated by five measures: three subdomains of the Unified Batten Disease Rating Scale (UBDRS) [29] and two neuropsychological assessments. The UBDRS is a clinical tool developed to quantitatively assess the signs and symptoms of CLN3 disease [29]. In this study, we focused on UBDRS scores from three specific subdomains: (a) Physical: assesses neurological impairment across 28 items on a scale from 0 (normal) to 4 (severely impaired), yielding a total score of 0–112, with higher scores indicate more severe impairment; (b) capability with actual vision: evaluates five activities of daily living on a scale of 1-14, where lower scores reflect greater functional impairment; (c) Clinical Global Impression (CGI): a provider rated measure of overall disease severity ranging from 6-35, with higher scores denoting greater severity [29]. The neuropsychological assessments included: (a) evaluation of adaptive functioning using the Vineland Adaptive Behavior, Third Edition (VABC-3), parental interview form [30]; and (b) assessment of verbal intelligence quotient (VIQ) using either the Wechsler Intelligence Scale for Children, Fourth Edition (WISC-IV) or the Wechsler Adult Intelligence Scale, Fourth Edition (WAIS-IV) [31, 32], both standardized with a mean of 100 and a standard deviation of 15, where higher scores reflect greater ability. Notably, UBDRS Physical and CGI are positively related to age and disease progression in CLN3, whereas UBDRS Capability, VABS-3 and VIQ scores are inversely related [33, 34].

Given the nature of the data described above and shown in Table 1, we split the dataset into two subsets: a classification subset and a severity subset (the two subsets can be found in a supplemental file named Supplementary_File_2). The classification subset composed of proteomics data collected via PEA or MS from both CLN3 (n=28) and non-CLN3 (n=45) participants. This subset was used to identify proteins that differentiate CLN3 from non-CLN3. The severity subset included proteomics data, clinical features derived from electrocardiogram (ECG) and vital signs, and laboratory tests, as well as UBDRS and neurodevelopmental assessment scores for all 42 CLN3 participants.

### 2.2 Data Imputation

In our dataset, approximately 32.3% of the proteomics measurements obtained through MS were missing, compared with 25% of the clinical measurements (i.e., ECG, vital signs and laboratory tests), and 16.4% of the disease severity scores. To create complete, unbiased, and representative datasets for predictive model development, we conducted data imputation on both subsets.

We evaluated multiple imputation methods, including PCA-based imputation [35], soft imputation [36], Random Forest (RF)-based imputation [37], and hot deck imputation [38]. To benchmark their performance, we introduced 10% pseudo-missing values into the dataset and compared the imputed values to the original true values. Evaluation metrics [39] included Mean Absolute Error (MAE) [40], Root Mean Square Error (RMSE) [41], Mean Square Error (MSE) [42], Raw Bias (RB), Percent Bias (PB), Coverage Rate (CR), and Average Width (AW) [43]. MAE and RMSE quantify the average and squared deviations with lower values indicating more accurate imputation. MSE, the square of RMSE, is more sensitive to large errors. RB measures systematic deviation and should ideally be near zero, while PB expresses this bias as a percentage, with <5% considered acceptable [39]. CR reflects the proportion of confidence intervals (CIs) that contain the true value (acceptable if ≥95%), and AW indicates the average CI width, with narrower widths denoting higher statistical efficiency. The formulas and detailed description for these evaluation metrics are listed in Table 2. The R packages *missMDA* [35], *softImpute* [36], *missforest* [37], *VIM* [44], *caret* [45], *foreach* [46], *doParallel* [47], *dplyr* [48], and *ggplot2* [49] were used for the imputation and performance evaluation.

**Table 2.**
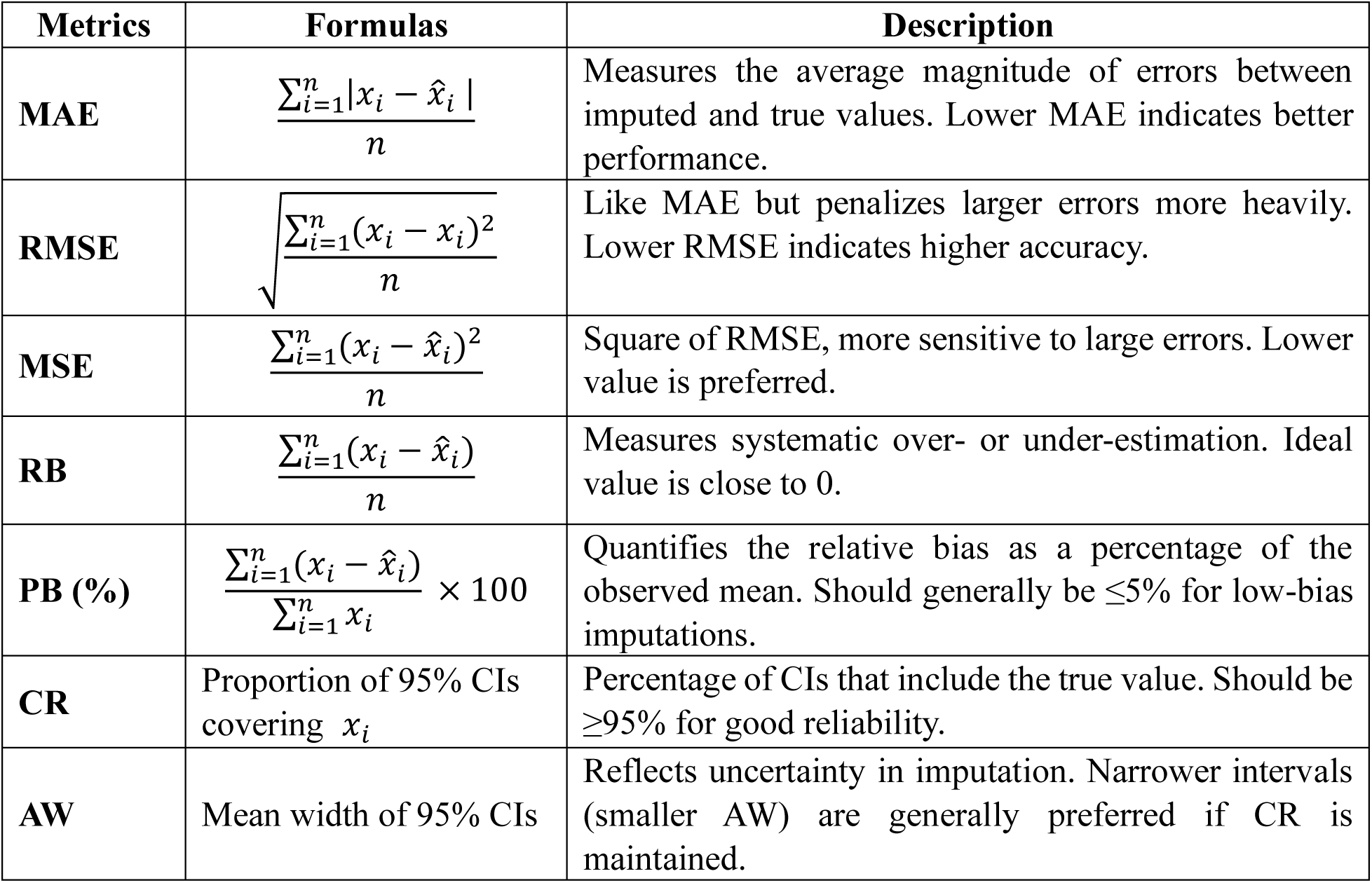
Imputation evaluation Metrics.

### 2.3 Predictive Model Development

To identify potential protein biomarkers associated with CLN3 disease and its severity, we developed multiple predictive machine learning models using the data summarized in Table 1 and evaluated their performances with various metrics.

#### 2.3.1 Model development with the classification subset

To build predictive models using the classification dataset for identifying features significantly associated with CLN3 disease, we implemented five algorithms: Logistic Regression [50] (with and without Least Absolute Shrinkage and Selection Operator (LASSO) regularization) [51], Random Forest (RF) [52], Support Vector Machines (SVM) [53], and eXtreme Gradient Boosting (XGBoost) [54]. All features were normalized using Z-score normalization [55] prior to modeling. The dataset was then randomly split into a training set (70%) and a testing set (30%) using stratified sampling via the createDataPartition() function from the *caret* package [45].

The baseline logistic regression model was developed using the glm() function [56] with the binomial family. For LASSO logistic regression, we employed the cv.glmnet() function from the *glmnet* package [57] to perform 10-fold cross-validation and identify the optimal regularization parameter (lambda.min). Final predictions were generated at the optimal lambda value. The RF model was trained using the *randomForest* package [58] with 500 trees. The SVM model was built using the svm() function from the *e1071* package [59], with probability estimation enabled. The XGBoost model was implemented using the *xgboost* package [60], configured with a binary logistic objective, a learning rate (eta) of 0.1, a maximum tree depth of 3, and 100 boosting rounds. Model performance was evaluated using multiple classification metrics, including the Area Under the Receiver Operating Characteristic Curve (AUROC), sensitivity, specificity, precision, F1 score, and balanced accuracy. These metrics were computed using a custom evaluation function built on the *caret* [45], *pROC* [61], and *dplyr* [48] packages. All model development and evaluation were conducted using the R programming environment.

#### 2.3.2 Model development with the severity subset

We developed six multivariate prediction models to identify features (i.e., proteins, clinical variables) associated with disease severity, as indicated by five severity scores. These models included Multivariate Linear Regression (MLR) [62], Partial Least Squares Regression (PLSR) [63], RF multivariate model [64], XGBoost Multi-Output Regression [54], Linear Regression with LASSO regularization [51], and Feedforward Neural Network (FNN) [65]. Prior to model training, all features were normalized using Z-score normalization. The dataset was split into a training set (70%) and a testing set (30%) using the train_test_split() function from the *scikit-learn* python library [66], with a fixed random seed to ensure reproducibility.

The MLR model was developed using the LinearRegression() class, while PLSR was implemented with the PLSRegression() function using five components. The RF model was trained with 500 trees using the RandomForestRegressor() function from *scikit-learn*. For XGBoost, independent regressors were built for each severity score using the XGBRegressor() class from the *xgboost* library [54], configured with a squared error objective, 100 boosting rounds, a learning rate of 0.1, and a maximum tree depth of 3. LASSO regression was performed using the MultiTaskLassoCV() function five-fold cross-validation. Lastly, an FNN model was implemented using the *Keras* API in the *TensorFlow* library [67]. The architecture included two hidden layers with 64 and 32 units (ReLU activation), followed by a linear output layer corresponding to the number of severity scores. The model was trained with the Adam optimizer for 100 epochs and a batch size of 16. Model performance was evaluated on each severity score using four key metrics: Mean Square Error (MSE), Root Mean Square Error (RMSE), Mean Absolute Error (MAE), and the coefficient of determination (R²) [68]. These metrics were calculated using functions from using the *scikit-learn* [66], *numpy* [69], *pandas* [70], and *matplotlib* [71] libraries. All model development using the severity dataset was conducted in Python.

### 2.4 Protein Candidate Identification

#### 2.4.1 Feature extraction from the classification subset

Feature extraction was performed using logistic regression with LASSO regularization, which demonstrated the best performance among the evaluated models (see Results section). We first identified the optimal regularization parameter (*α*) by training LASSO logistic regression models over a range of *α* value (0 to 1) using 5-fold cross-validation. The model with the highest AUROC was selected for further analysis. To improve the robustness of the feature selection process, we employed a bootstrapped stability selection approach. Specifically, 200 bootstrap samples were generated by randomly resampling participants with replacement from the original dataset. For each bootstrap iteration, a LASSO logistic regression model was fitted using the previously identified optimal *α* and the selected features were recorded. We then calculated the selection frequency for each feature across all 200 bootstrap models. Features appearing in at least 80% of bootstrap iterations were selected and retained for subsequent analysis. All modelling and feature section were implemented in R using the *glmnet* [57] and *caret* packages.

#### 2.4.2 Feature extraction from the severity subset

The multivariate RF regression model, which achieved the superior performance within the severity subset, was selected to identify features significantly associated with the five disease severity scores. For each disease severity score, we trained RF models on 50 bootstrap samples generated by randomly resampling the original dataset with replacement. Feature importance was assessed using tree-based metrics, particularly mean minimal depth, which measures how early a feature is used in tree splits, with smaller values indicating greater predictive influence. To determine robust and influential feature selection, importance scores were normalized via Z-score normalization. Features with normalized importance scores greater than 2 were selected and retained as stable predictive features for downstream analysis. All analyses were conducted in R using the *randomForest* package [58] and the *randomForestExplainer* package [72].

#### 2.4.3 Biological Interpretation through pathway enrichment analysis

To interpret the biological relevance of the identified protein features and describe their underlying functional context, we performed pathway enrichment analysis on them. These features identified from both the classification and severity subsets were first merged, and unique features were retained for analysis. These features were stratified into upregulated or downregulated groups based on their differential expression in CLN3 participants relative to controls. Each group was analyzed independently to uncover biological pathways significantly enriched in the disease state. The analysis was conducted using the Metascape platform [73] using the curated gene sets from the Kyoto Encyclopedia of Genes and Genomes (KEGG) [74], Reactome [75], and Gene Ontology (GO) biological processes [76]. Enrichment results were filtered using stringent criteria:

Benjamini-Hochberg adjusted p-value < 0.01 and enrichment fold > 1.5 [73] to ensure statistical robustness and biological relevance.

### 2.5 Protein Candidate Prioritization via Protein-Protein Interaction (PPI) Network

To prioritize the identified protein features with potential regulatory roles, we performed network-based analysis within a protein-protein interaction (PPI) network by assessing their topological importance. All proteins were queried in the STRING database [77] to construct a high-confidence and the resulting network was analyzed using Cytoscape [78].

To evaluate the topological importance of each protein, we applied five complementary centrality measures using CytoHubba plugin (v0.1) [79], which quantify a node’s influence within the PPI.

- Degree Centrality (DC): Number of direct connections to a node (i.e., protein) [80];
- Betweenness Centrality (BC): Percentage of shortest paths passing through a node (i.e., protein) [81];
- Closeness Centrality (CC): Inverse of the average shortest path of the node (i.e., protein) to all other nodes [82];
- Maximum Clique Centrality (MCC): Membership in densely connected subnetworks [83];
- Degree of Maximum Neighborhood Component (DMNC): Importance based on local neighborhood connectivity [83].

To generate a consensus ranking, we normalized each centrality score to a 0–1 scale and computed an overall centrality score by averaging the normalized values across all five metrics. Proteins were ranked based on this aggregated score, and the top 20 proteins were selected as candidate biomarkers for further analysis. Additionally, we visualized the top 10 proteins from each centrality measure using a multi-line dot plot to highlight consistent hub proteins and centrality score distributions across the metrics.

### 2.6 Corroboration of Candidate Biomarkers using External CLN3 Omics Data

To independently corroborate the diagnostic potential of the identified candidate biomarkers, we performed external corroboration using a publicly available transcriptomic dataset (GEO accession: GSE22225) [84], obtained from the Gene Expression Omnibus (GEO) repository [85].

This dataset includes lymphocyte gene expression profiles of 8 CLN3 patients with variable disease progression and 7 healthy controls.

#### 2.6.1 Data processing and gene mapping

Raw microarray data files (.CEL format) were preprocessed using the Robust Multi-array Average (RMA) algorithm, implemented via the *affy* R package (v1.84.0). After normalization and summarization of gene-level expression values, we extracted the expression profiles corresponding to the top proteins identified from the above step. Protein-to-gene mapping was conducted using UniProt and platform-specific annotations to ensure accurate correspondence between proteomic features and transcriptomic probes.

#### 2.6.2 Diagnostic evaluation of candidate biomarkers

To assess the discriminative power of these candidate biomarkers, we performed receiver operating characteristic (ROC) curve analysis for each gene corresponding to the top 20 proteins. AUROC (Area Under the ROC Curve) values were calculated to quantify the classification performance of each gene in distinguishing CLN3 patients from healthy controls. Based on AUROC values, the six genes with the highest discriminative accuracy were selected as the most promising diagnostic biomarkers in this independent cohort.

## 3. Results

### 3.1 Data Imputation Performance Evaluation

To determine the most suitable imputation method for our datasets, we compared four imputation methods using various evaluation metrics (Table 3) and performance of these imputation methods by density comparison and scatter plot can be found in the Supplementary_Figure_1. We used MAE and RMSE to assess primary performance, as they measure the average and squared deviations between full and imputed values, key indicators of predictive reliability [86, 87]. Methods with lower MAE and RMSE values indicates better accuracy between the imputed and true values. Secondary metrics included PB, RB, CR, and AW, to evaluate statistical fidelity, distribution preservation, and interval coverage [43]. This multi-metric evaluation ensured that the selected imputation methods maintained both model accuracy and data integrity.

**Table 3.**
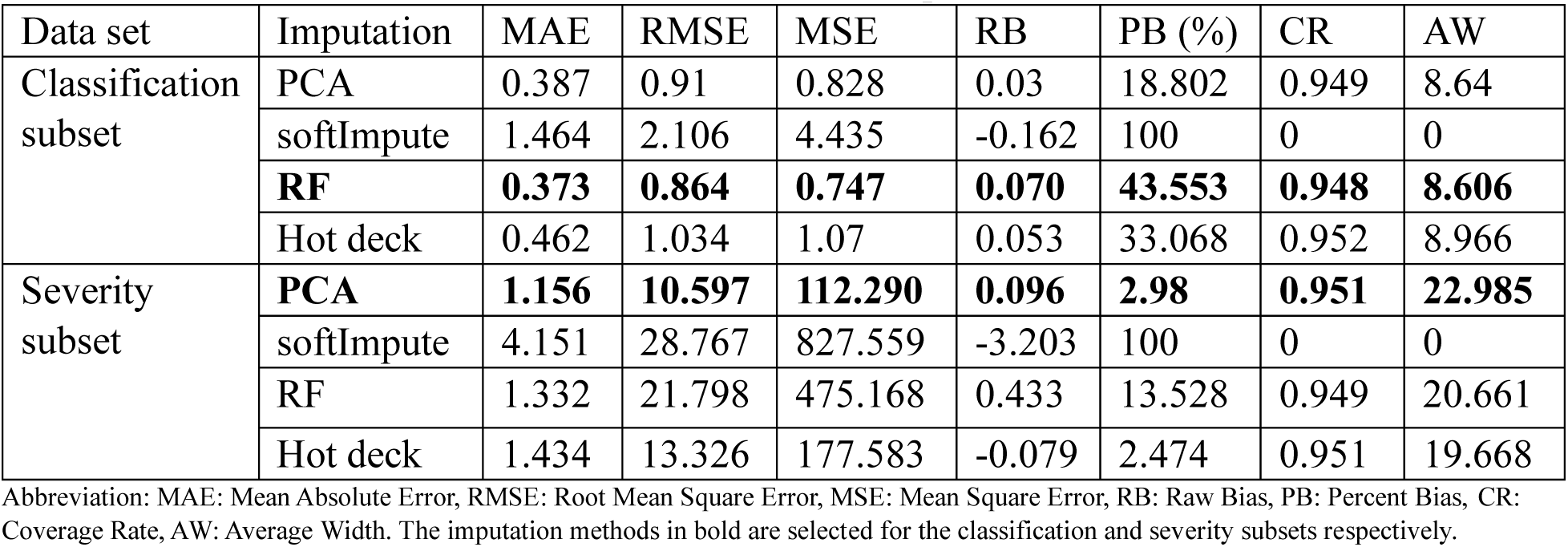
Evaluation Metrics for Imputation Methods.

#### 3.1.1 Classification subset

Among the evaluated methods, RF-based imputation performed best for the classification dataset, yielding the lowest MAE (0.373) and RMSE (0.864). PCA-based imputation was a close second, exhibiting a slightly higher MAE (0.387) but a lower percent bias (PB = 18.8%), suggesting minimal systematic bias. Although hot deck imputation preserved the highest coverage rate (CR = 0.952), its elevated error rates (MAE = 0.462 and RMSE = 1.034) limited its utility. Based on the overall performance, RF-based imputation was selected for the classification dataset.

#### 3.1.2 Severity subset

For the severity dataset, PCA-based imputation demonstrated superior performance, achieving the lowest MAE (1.156), RMSE (10.597), and PB (2.98%), indicating high accuracy and low bias. While hot deck imputation again preserved distribution well (CR = 0.951), its higher error values (MAE = 1.434 and RMSE = 13.326) made it less suitable. RF-based imputation showed moderate performance but was inferior in both predictive error and bias. Consequently, PCA-based imputation was selected for the severity subset.

### 3.2 Prediction Model Development and Feature Identification

#### 3.2.1 Classification models for identifying CLN3 diagnostic proteins

To identify the most effective classification approach for distinguishing CLN3 patients from non-CLN3 participants based on proteomic profiles, we developed and evaluated five classification models. Among these, LASSO logistic regression outperformed all others across key evaluation metrics (Table 4), achieving the highest accuracy (0.905), AUROC (0.885), and balanced accuracy (0.923). It also demonstrated perfect sensitivity (1.00) and high specificity (0.846), indicating strong discriminative power with a low false positive rate and no false negatives.

**Table 4.** Performance evaluation for classification models.

| Models | Accuracy | Sensitivity | Specificity | Precision | F1 | Balanced Accuracy | AUROC |
| --- | --- | --- | --- | --- | --- | --- | --- |
| Logistic | 0.524 | 0.75 | 0.384 | 0.429 | 0.545 | 0.567 | 0.567 |
| **LASSO** | **0.905** | **1** | **0.846** | **0.8** | **0.889** | **0.923** | **0.885** |
| RF | 0.714 | 0.375 | 0.923 | 0.75 | 0.5 | 0.649 | 0.837 |
| SVM | 0.667 | 0.5 | 0.769 | 0.571 | 0.533 | 0.635 | 0.846 |
| XGBoost | 0.762 | 0.5 | 0.923 | 0.8 | 0.615 | 0.712 | 0.837 |

Abbreviations: RF= Random Forest, SVM= Support Vector Machine, AUROC= Area Under the Receiver Operating Characteristic Curve. The classification model in bold shows superior performance.

In contrast, standard logistic regression exhibited poor performance, particularly in specificity (0.384), indicating a high false positive rate. While the RF model achieved high specificity (0.923), its sensitivity was markedly low (0.375), limiting its diagnostic utility due to frequent false negatives. SVM and XGBoost showed intermediate performance, both with a sensitivity of 0.5; their F1 scores were 0.533 and 0.615, respectively, but neither matched the balanced and robust performance of the LASSO model. Based on these results, LASSO logistic regression was selected as the optimal classifier for identifying diagnostic protein biomarkers for CLN3.

#### 3.2.2 Prediction models for identifying protein associated with CLN3 disease severity

To identify proteins significantly associated with CLN3 disease severity, we developed and evaluated six predictive models using four performance metrics: RMSE, MAE, R², and adjusted R² (Table 5). Among these models, RF outperformed all others, achieving the lowest RMSE (7.474) and MAE (6.583) as well as the highest R² (0.301). These results suggest that RF can capture non-linear relationships and complex feature interactions, which is particularly well-suited for modeling the capturing heterogeneity observed in CLN3. Further analysis of individual disease severity scores (Supplementary_File_3) reinforced the robustness of the RF model, which yielded the highest predictive accuracy for UBDRS Physical Weighted Score (R² = 0.273), Capability Weighted Score (R² = 0.324), and CGI Weighted Score (R² = 0.291). In contrast, MLR and PLSR performed poorly, with negative R² values, indicating poor model fit. LASSO and XGBoost showed modest improvement but remained suboptimal overall. The FNN model exhibited signs of overfitting or convergence failure, characterized by a highly negative R² (−14.818) and inflated RMSE, making it unsuitable for this dataset. Full results of the evaluation metrics for the models applied to the severity dataset can be found in a supplemental file named Supplementary_File_3.

**Table 5.**
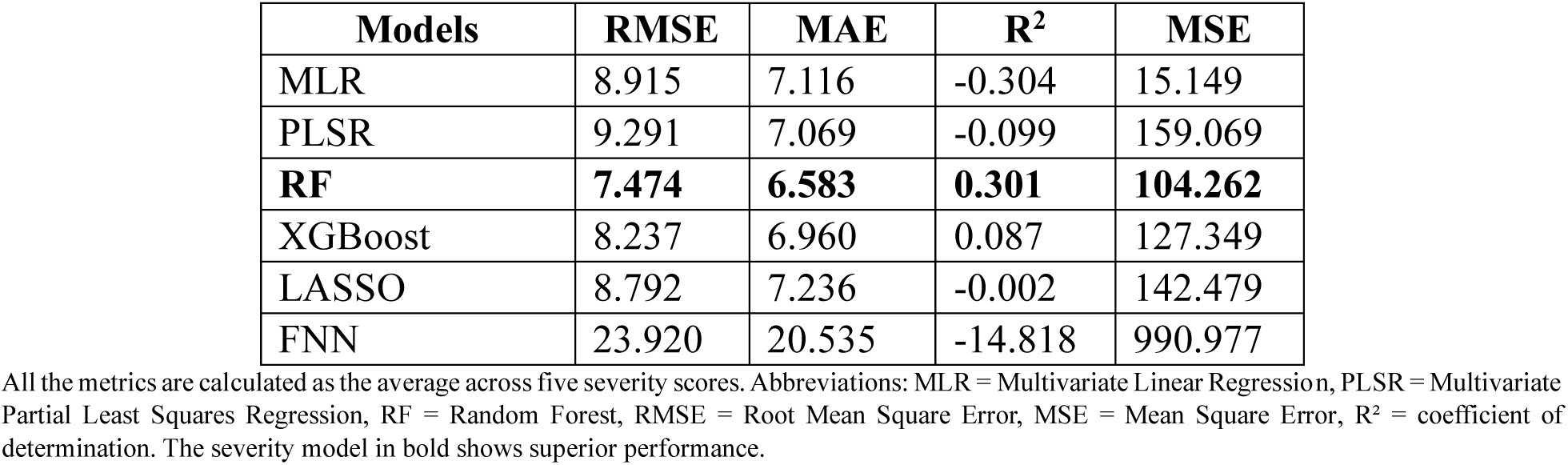
Performance evaluation of severity models.

| **Models** | **RMSE** | **MAE** | **R<sup>2</sup>** | **MSE** |
| --- | --- | --- | --- | --- |
| MLR | 8.915 | 7.116 | -0.304 | 15.149 |
| PLSR | 9.291 | 7.069 | -0.099 | 159.069 |
| **RF** | **7.474** | **6.583** | **0.301** | **104.262** |
| XGBoost | 8.237 | 6.960 | 0.087 | 127.349 |
| LASSO | 8.792 | 7.236 | -0.002 | 142.479 |
| FNN | 23.920 | 20.535 | -14.818 | 990.977 |

All the metrics are calculated as the average across five severity scores. Abbreviations: MLR = Multivariate Linear Regression, PLSR = Multivariate Partial Least Squares Regression, RF = Random Forest, RMSE = Root Mean Square Error, MSE = Mean Square Error, R<sup>2</sup> = coefficient of determination. The severity model in bold shows superior performance.

Based on its superior and consistent performance across multiple severity components, the RF model was selected as the optimal model for severity prediction and was subsequently used to identify candidate protein biomarkers.

#### 3.2.3 Feature identification

The optimized LASSO classification model identified 32 proteins, while the RF severity model identified 234 features. After merging the results and removing duplicates, 260 unique proteins and three metabolites [3-OH-decenoylcarnitine (C10:1-OH), 3-OH-hexanoylcarnitine (C6-OH), and Formiminoglutamate (FIGLU)] were retained for further analysis. A complete list of identified features is provided in the Supplementary_File_4. As the primary goal of this study is to identify diagnostic biomarkers, only protein candidates were applied in the following steps. We would further analyze other features in the near future.

#### 3.2.4 Biological Interpretation through functional Enrichment Analysis

To contextualize the biological relevance of the proteins identified through the classification and severity prediction models, we performed pathway enrichment analysis using gene sets from KEGG, Reactome and GO. The analysis revealed widespread dysregulation of multiple biological processes in CLN3, involving both upregulated and downregulated proteins. Significantly enriched pathways (Benjamini-Hochberg adjusted *p* < 0.01 and enrichment fold > 1.5) are presented in Figure 2.

**Figure 2.**
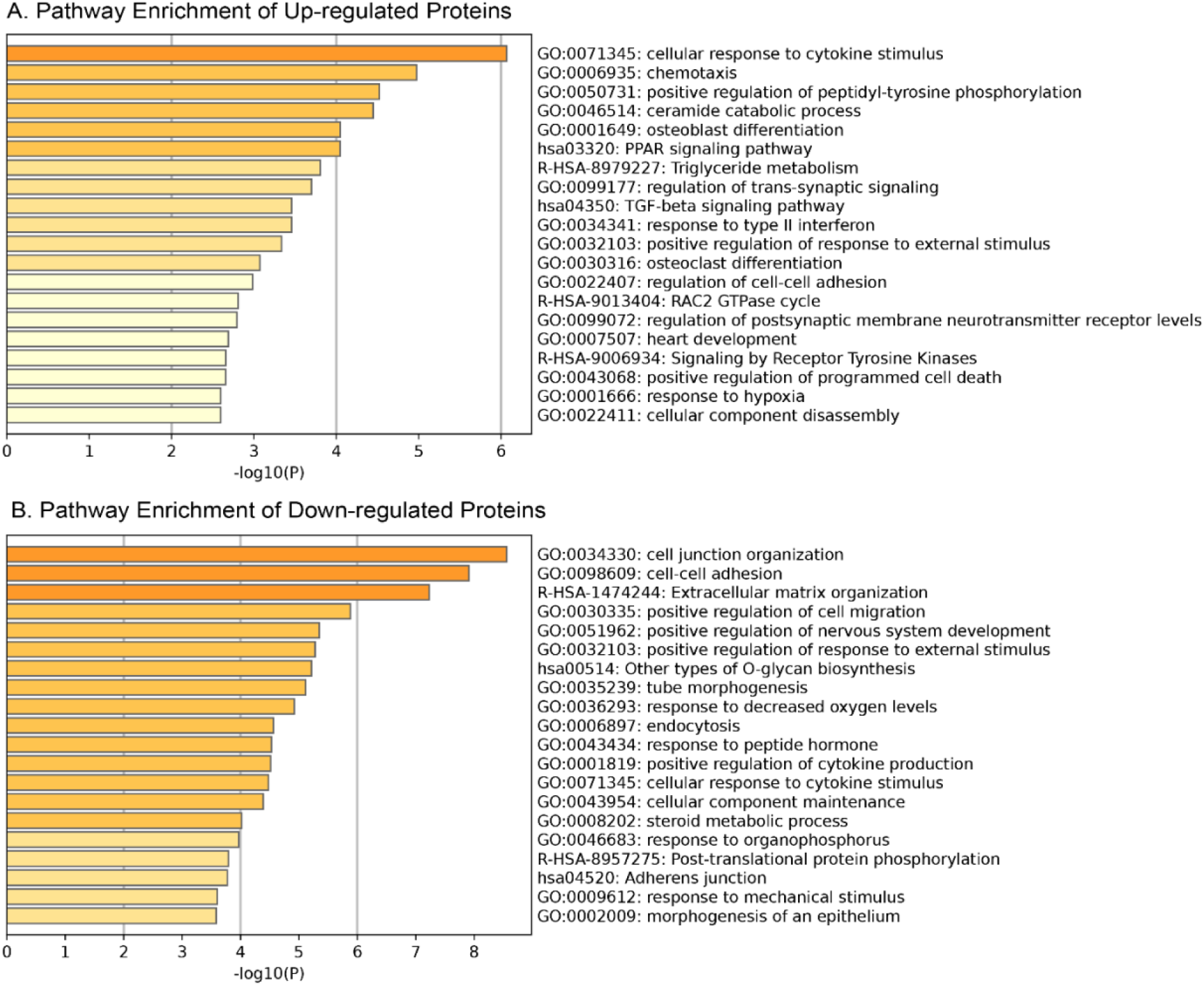
Top enriched pathways of the up/down regulated proteins.

As shown in Figure 2-A, upregulated proteins were strongly enriched in pathways related to immune activation and inflammatory responses, including cellular response to cytokine stimulus, chemotaxis, TGF-beta signaling, and type II interferon [88–90]. In addition, several metabolic processes were overrepresented, such as, ceramide catabolic process, PPAR signaling, and triglyceride metabolism [3, 4, 91]. Neurological processes including regulation of trans-synaptic signaling and postsynaptic neurotransmitter receptor levels [92, 93] were also elevated, alongside stress response pathways like regulation of programmed cell death and response to hypoxia [94, 95]. Further enrichment was observed in receptor tyrosine kinase signaling and RAC2 GTPase cycle [96, 97], implicating dysregulated cell signaling and adhesion.

Conversely, as shown in Figure 2-B, downregulated proteins were predominantly enriched in pathways associated with structural integrity and intercellular communication. These included cell junction organization, cell-cell adhesion, adherent junctions, and extracellular matrix organization [98, 99]. Additional suppressed pathways were associated with cell migration, epithelial and tube morphogenesis [4, 100], and nervous system development [5], further highlighting disruptions in key developmental processes. Metabolic and homeostatic deficits were also evident, reflected by reduced enrichment in pathways related to endocytosis, O-glycan biosynthesis, cellular responses to peptide hormone and mechanical stimulus [5, 101–104].

Collectively, these results illustrate a dual pathologic landscape in CLN3: on one hand, heightened immune activation and neurodegenerative signaling, and on the other, suppressed cellular architecture, developmental pathways, and homeostatic regulation. This biological context not only enhances the interpretability of the identified biomarkers but also provides mechanistic insight for future therapeutic strategies.

### 3.2 PPI Network-Based Protein Prioritization

The PPI network was constructed by querying the 260 identified unique proteins via the STRING database and visualized in Cytoscape. The resulting network consists of 258 nodes and 393 edges, representing direct physical interactions as well as functional associations. Two proteins, one pseudogene (AHCYP2), and one annotation derived duplicate were excluded from downstream analysis. Proteins were ranked based on a composite centrality score, calculated as the mean of normalized values across all five metrics. This analysis resulted in 20 unique proteins with the highest network centrality: EGFR, HIF1A, CXCL12, CSF1, VEGFC, COL4A3, VEGFD, ERBB4, OSM, NPM1, CD74, BRD4, TFRC, STAG2, LMNB1, MYH9, IL6R, COLGALT2, COL6A1, and CTCF. There proteins were considered as potential biomarker candidates for further study.

Figure 3 shows the top 10 proteins ranked by each centrality measure. Several proteins including EGFR and HIF1A, consistently scored highly across five centrality metrics, suggesting their critical role as central regulators in the CLN3-associated molecular network. To better contextualize the network topology of key proteins, we constructed a core subnetwork centered on Closeness Centrality (CC), a metric that prioritizes nodes with short average path to all other. This subnetwork included the top 20 proteins ranked by CC, their immediate first-degree interactors, and the shortest paths connecting them. As shown in Figure 4, this subnetwork demonstrates dense interconnectivity among central nodes, indicating that these hub proteins may coordinate multiple biological pathways related to CLN3 pathogenesis. These findings support the biological plausibility of those hub proteins as candidate biomarkers or therapeutic targets, particularly those with both high centrality and functional relevance in enrichment analyses.

**Figure 3.**
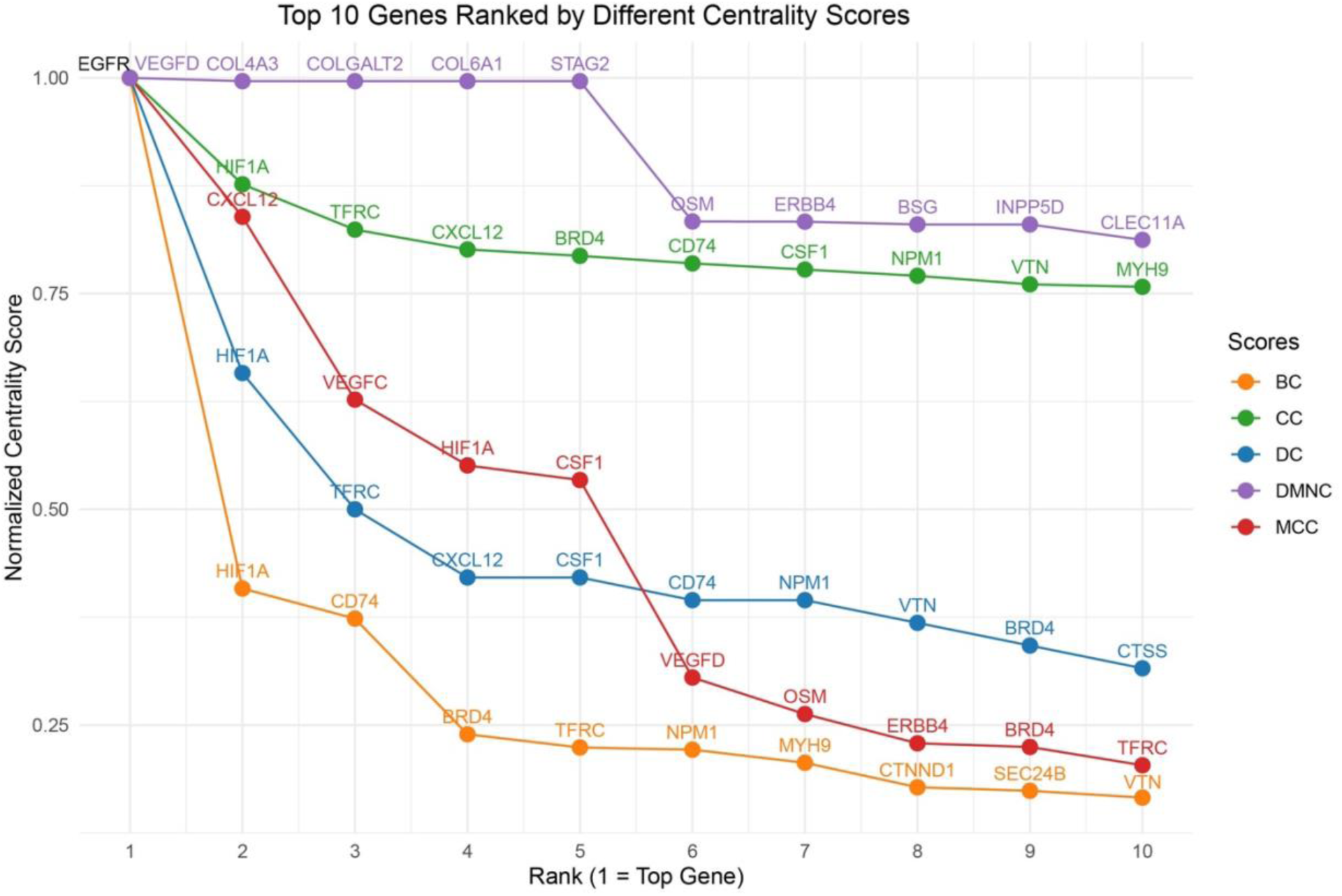
Top 10 proteins ranked by five centrality measures in the CLN3-associated PPI network. Five centrality scores were normalized to a 0–1 range for comparability. Each line represents one centrality metric, with the x-axis indicating rank position (1–10) and the y-axis showing normalized centrality scores. Overlap of top-ranked proteins across metrics (such as EGFR and HIF1A) suggests robust hub roles and potential functional importance in CLN3-related pathways. Abbreviations: DC = Degree Centrality; BC = Betweenness Centrality; CC = Closeness Centrality; MCC = Maximum Clique Centrality; DMNC = Degree of Maximum Neighborhood Component.

**Figure 4.**
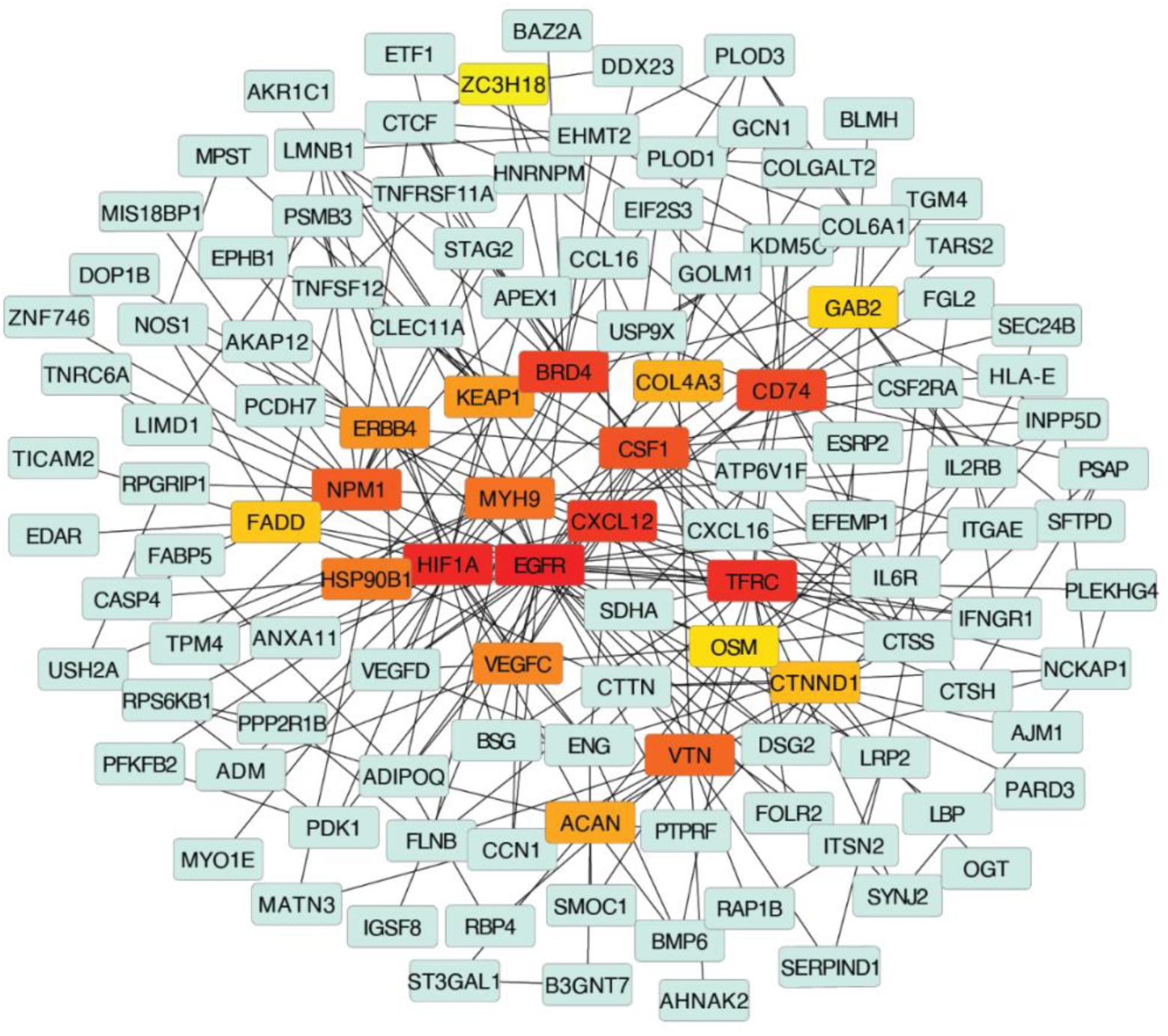
Subnetwork with Top 20 Hub Genes. Nodes colored from red to yellow represent the top 20 hub proteins ranked by the Closeness Centrality (CC). The red indicates the highest rank while the yellow indicates the lowest rank. Green nodes indicate proteins that directly interact with these top hub proteins.

### 3.4 Corroboration of Candidate Biomarkers with an External CLN3 Patient Dataset

Using an independent GEO dataset GSE22225, we observed six proteins among 20 identified candidate proteins (OSM, IL6R, LMNB1, HIF1A, NPM1 and CSF1) exhibiting strong discriminatory power, each achieving an AUROC greater than 0.8 (Figure 5). Violin plots further illustrate these genes expression differences between CLN3 and non-CLN3 samples (Figure 6). As the clinical courses CLN3 patients were classified into three groups: slow, average or rapid disease progression defined by the Index of Relative Severity [105]. We then examined the relative expression among CLN3 patients with different progression rates. The results showed that several of the top proteins through centrality analysis, such as OSM and HIF1A, demonstrated marked differential expression in CLN3, particularly among those with slow disease progression. Additionally, the expression level of gene LMNB1 was progressively elevated in subgroups with faster disease progression, suggesting their potential utility as prognostic biomarkers (Figure 7). These findings support the robustness of our biomarker selection and indicate that these six genes, individually and potentially in combination, may serve as effective diagnostic markers for CLN3 disease in clinical samples. ROC curves and expression plots for all 20 proteins are provided in Supplementary_Figure_2.

**Figure 5.**
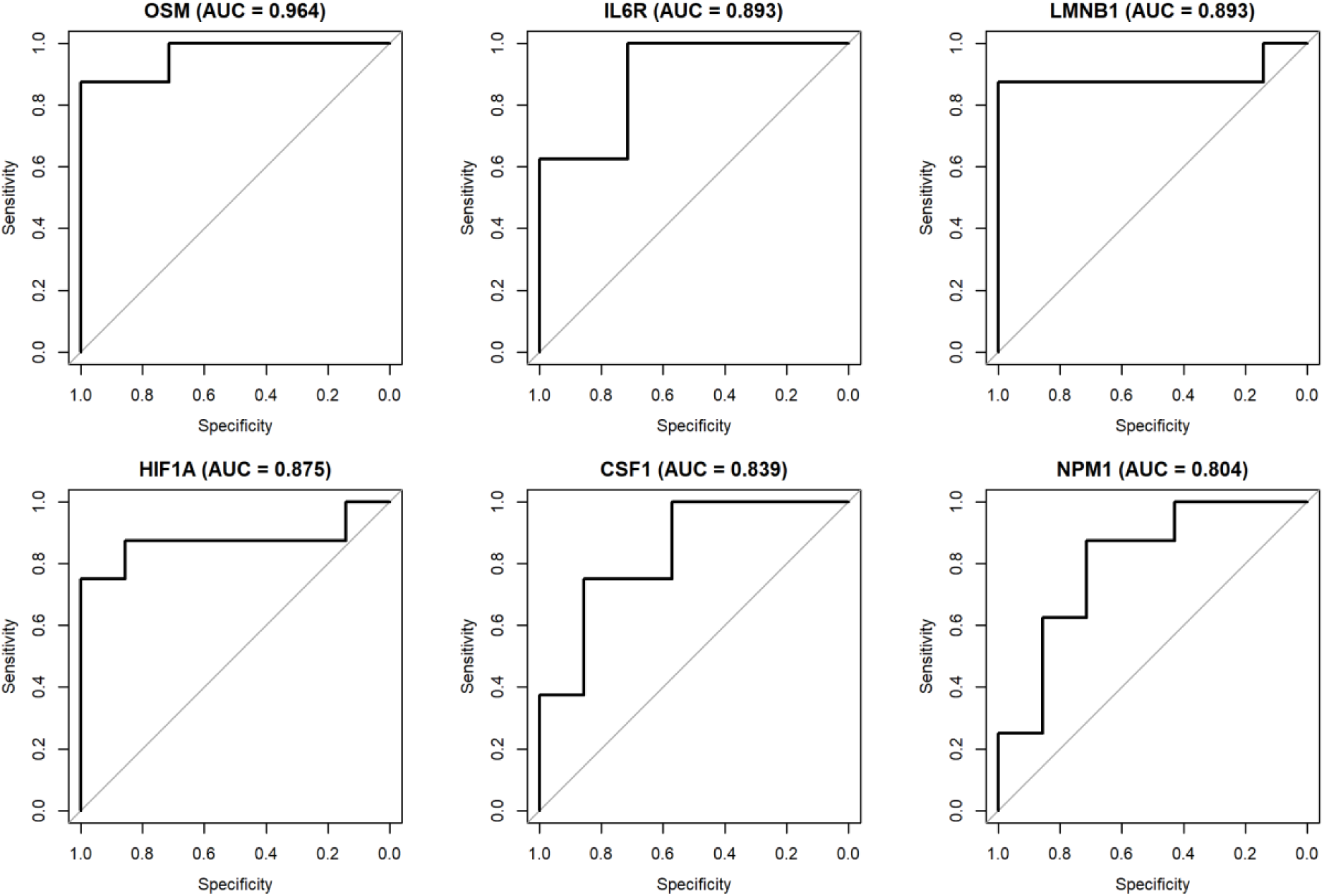
Top six proteins with strong discriminatory power (AUC > 0.8)

**Figure 6.**
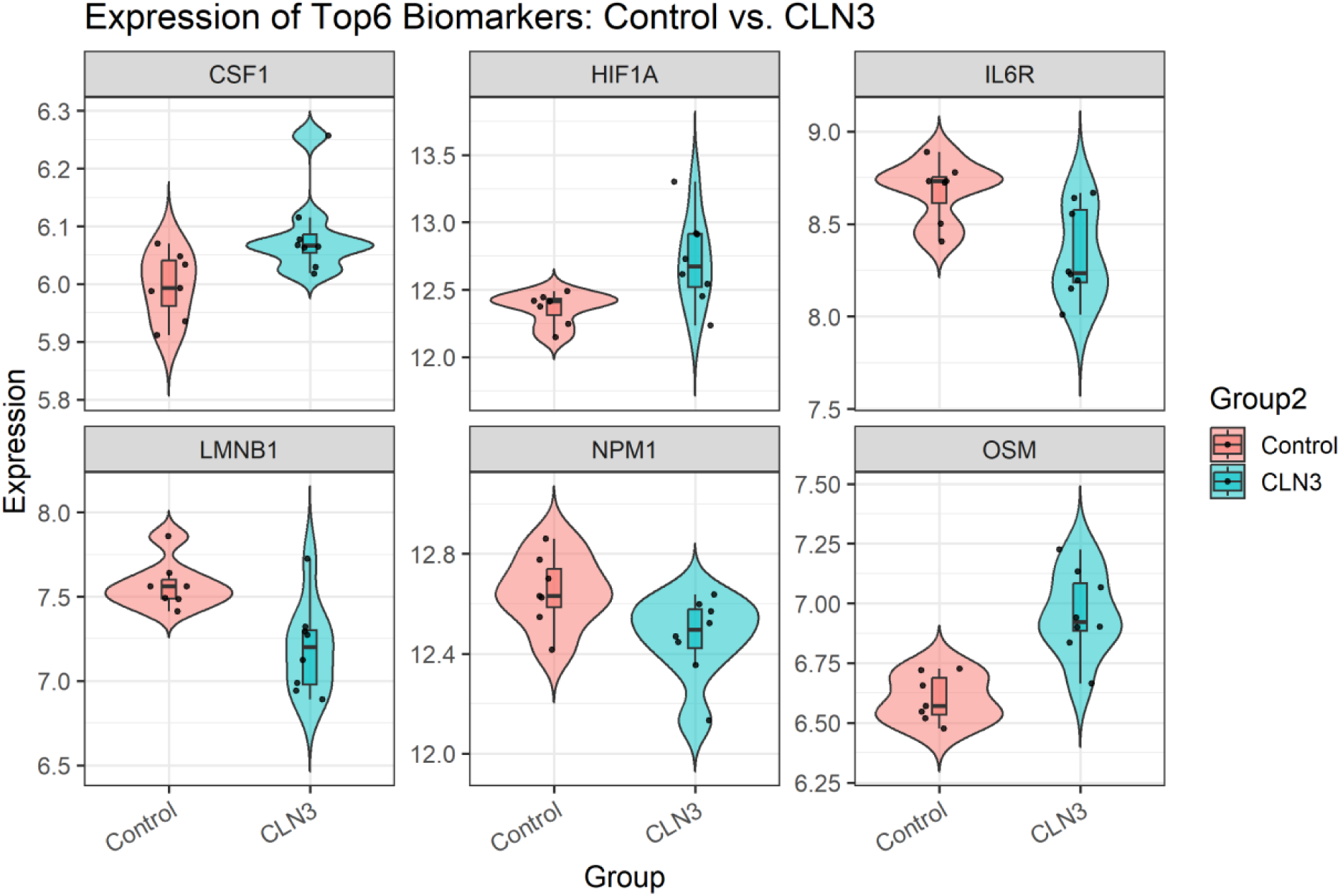
Differential Expression of Top 6 Biomarkers in Control vs. CLN3 Groups.

**Figure 7.**
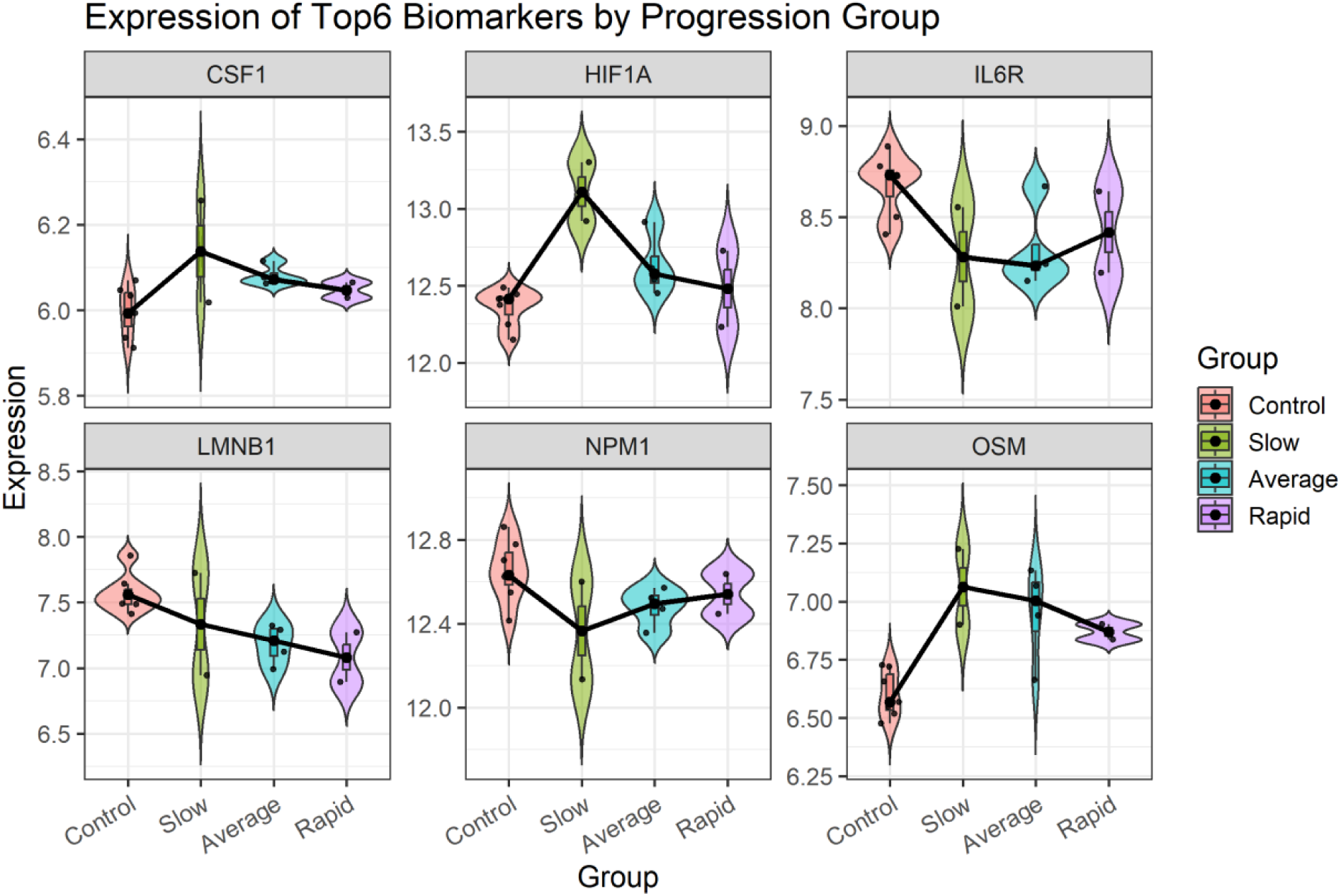
Differential expressions of the top six biomarkers in control and CLN3 patient groups stratified by disease progression speed.

### 3.5 Mechanism Insights of Biomarker Candidates

OSM (Oncostatin M) emerged as the most promising candidate. OSM is a multifunctional cytokine involved in inflammatory signaling and tissue remodeling [106]. Its receptor, OSMR, has shown differential protein expression in CLN3 patients and was closely related to biomarker candidates in a previous study [15]. OSM impacts neurodevelopment and neuroinflammation via STAT3-mediated pathways, affecting astrocytic glutamate uptake and blood-brain barrier integrity, while also contributing to neuroprotection and remyelination following neural injury [106–108]. Increased OSM expression might exacerbate neuroinflammatory responses and disrupt glial-neuronal homeostasis in CLN3, potentially influencing disease progression.

HIF1A (Hypoxia-Inducible Factor 1 Alpha) is a transcription factor that coordinates cellular responses to hypoxia and inflammation [109]. While not directly studied in CLN3, research in other lysosomal storage disorders, such as Niemann-Pick disease type C1 (NPC1), demonstrates that HIF1A-mediated pathways modulate neuroinflammation and cellular stress [110, 111]. Interactions between HIF1A, immune activation, and lysosomal dysfunction may influence the neurodegenerative processes observed in CLN3 disease.

LMNB1 encodes lamin B1, a key structural protein of the nuclear lamina that is essential for maintaining nuclear envelope integrity and regulating neurodevelopmental processes, including neuronal differentiation, migration, and survival [112]. In silico analysis of an external CLN3 dataset showed that LMNB1 expression was negatively correlated with the speed of CLN3 disease progression, suggesting that reduced LMNB1 may impair neuronal migration and survival, contributing to disease severity. Previous studies also indicate that LMNB1 deficiency disrupts cortical layering and may contribute to neurodevelopmental abnormalities in CLN3 [112–114].

IL6R (Interleukin-6 Receptor) mediates signal transduction through the JAK/STAT pathway and is critical for inflammatory and immune regulation [115]. It has shown differential expression in CLN3 patients and was closely related with biomarker candidates in a previous study [15]. Although direct evidence in CLN3 is limited, studies indicate CLN3 is required for proper JAK/STAT signaling during tissue regeneration [116], suggesting a possible interaction between IL6R activity and CLN3 function, with implications for immune dysregulation in CLN3 pathology.

NPM1 (Nucleophosmin 1) is involved in protein homeostasis, nucleolar function, and cellular stress responses [117]. Literature suggests that NPM1 may interact with CLN3 mRNA and influence stress granule formation [118]. While direct links to CLN3 neurodegeneration remain to be established, decreased NPM1’s activity affecting stress signaling aligns with cellular deficits seen in CLN3 pathology.

CSF1 (Colony Stimulating Factor 1) and its receptor, CSF1R, regulate the differentiation and survival of microglia and macrophages [119]. Altered CSF1-dependent glial responses have been implicated in neuroinflammatory mechanisms contributing to CLN3 progression [119–121]. Although direct causative evidence is limited, increased innate immune dynamics mediated by CSF1 likely play a role in the neuroinflammation of CLN3 disease.

In addition to proteins, three metabolites, 3-OH-decenoylcarnitine (C10:1-OH), 3-OH-hexanoylcarnitine (C6-OH), and formiminoglutamate (FIGLU), were identified as biomarker candidates. Elevated levels of C10:1-OH and C6-OH, which are hydroxylated fatty acids, are associated with disruptions in fatty acid β-oxidation and mitochondrial dysfunction [122, 123], processes which are implicated in CLN3 pathology [124, 125]. Formiminoglutamate (FIGLU) is a marker of folate metabolism, and disturbances in folate pathways have been linked to neurodevelopmental and neurodegenerative disorders such as Alzheimer disease and amyotrophic lateral sclerosis [126]. Although direct evidence of these metabolites in CLN3 is limited, their identification as predictive features suggests that metabolic dysregulation may contribute to disease progression and may serve as accessible biomarkers for monitoring CLN3.

## 4. Discussion

In this study, we systematically identified 263 biomarker candidates for CLN3 disease by analyzing the proteomics and clinical data pertinent to CLN3, ultimately prioritizing six proteins as the most promising candidates, i.e., OSM, IL6R, LMNB1, HIF1A, NPM1, and CSF1. These genes are involved in key processes such as brain development, immune response, and cellular stress management, and may contribute to nerve cell damage and brain inflammation observed in CLN3. Notably, LMNB1 was negatively correlated with disease progression speed, suggesting potential prognostic value, while literature supports the involvement of the other candidates in relevant pathological mechanisms associated with CLN3. Our findings highlight the effectiveness of the computational strategy we applied here for biomarker discovery for CLN3 and underscore its potential for broader application to other rare diseases and related conditions.

In this study, we introduced a computational framework composed of three components, 1) machine learning models with optimized imputation methods to analyze proteomics and laboratory data from CLN3 patients to identify protein candidates; 2) PPI network-based network analysis to prioritize candidates; and 3) corroboration of protein candidates using external gene expression datasets. This framework offers several advantages, first, the use of optimized imputation methods reduced bias and improved the reliability of the dataset for downstream modeling; second, selecting the best-performing models and incorporating bootstrapping enhanced robustness and generalizability of the models; third, network analysis enabled the prioritization of proteins with potentially pivotal biological roles; and finally, external corroboration confirmed six final biomarker candidates. Despite these strengths, several limitations remain. In this study, the proximity extension assay (PEA) was used to capture only a predesigned panel of approximately 1,500 proteins, limiting proteome-wide discovery. Additionally, the relatively high proportion of missing values and small sample sizes introduce risks of bias during imputation and model training, underscoring the need for more curated and comprehensive datasets in rare disease research. While multiple machine learning algorithms were evaluated, challenges related to data sparsity and heterogeneity persist. For instance, the poor performance of the feedforward neural network (FNN) model highlights the difficulty of applying deep learning approaches in small-scale datasets and points to the need for more advanced, sample-efficient deep learning methods in future work.

The two proteomics datasets analyzed in this study have previously been used for biomarker discovery efforts [15, 18]. In contrast to earlier analyses that primarily focused on differential expression and correlation-based approaches, we re-analyzed these datasets by integrating them with additional clinical data and applying a predictive, machine learning–driven framework. Our approach prioritized protein candidates based on their ability to predict disease classification and severity, rather than solely on statistical significance of expression differences. Notably, among the top 20 proteins ranked through PPI network–based centrality analysis, IL6R and VEGFD overlapped with previously reported differentially expressed proteins. This partial overlap highlights a key methodological distinction: our computational approach allowed us to uncover protein biomarkers with high predictive value that may not exhibit the strongest differential signals, thereby complementing conventional statistical approaches. In the meantime, we acknowledge that external corroboration was restricted by the limited availability of suitable datasets, which prevented more comprehensive analyses such as survival analysis or focused exploration of immune and transcription factor genes. The absence of longitudinal patient data also limits the robust assessment of prognostic biomarkers like LMNB1; thus, future studies should prioritize the collection of longitudinal CLN3 data to strengthen biomarker validation and potentially identify additional candidates predictive of disease progression.

To further elucidate the molecular mechanisms underlying CLN3 pathology, we performed functional enrichment analysis on the identified protein features. The enrichment results revealed pronounced upregulation of immune and inflammatory pathways, aligning with prior studies that implicate chronic neuroinflammation as a hallmark of CLN3 [88–90]. Enrichment of metabolic pathways, particularly those involved in ceramide catabolism and PPAR signaling, further emphasizes the critical roles of lysosomal dysfunction and disrupted lipid metabolism in disease progression [3, 4, 91]. Additionally elevated signaling related to synaptic function and programmed cell death corresponds with established evidence of neurodegeneration and synaptic dysfunction in CLN3, reflecting increased neuronal vulnerability and impaired neurotransmission [92–95]. Conversely, downregulated pathways related to cell adhesion, extracellular matrix organization, and endocytosis suggest deteriorating tissue architecture and compromised cellular maintenance [4, 5, 98–104, 127]. Notably, several dysregulated pathways involved in lipid metabolism and membrane remodeling reinforce the central pathogenic role of aberrant lipid storage and turnover in CLN3 disease [3, 91, 102, 128–131]. The results are inherently constrained by the scope and completeness of the functional annotation databases used (KEGG, Reactome, and GO), which may not fully capture the complexity or specificity of CLN3-specific molecular mechanisms. Furthermore, pathway annotations are often biased toward well-characterized diseases and may overlook rare disease–specific interactions. Neverthelss, these findings present a comprehensive view of CLN3 pathology, highlighting the interconnected roles of neuroinflammation, metabolic dysregulation, synaptic dysfunction, and impaired tissue architecture. These results underscore the biological relevance of the identified biomarker candidates and support their potential utility in advancing our understanding of CLN3 disease mechanisms.

## 5. Conclusion

This study demonstrates the effectiveness of the computational biomarker discovery framework in rare diseases and provides novel insights into the underlying molecular mechanisms of CLN3. The identification of new biomarker candidates, including proteins and metabolites, lays the groundwork for future experimental validation and the development of improved diagnostic and prognostic tools for CLN3 and potentially other rare diseases.

## Acknowledgement

We value the continued motivation and inspiration provided by study participants and their families and appreciate their dedication to help move research forward. This study was supported by funding from NIH intramural research (ZIA TR000548 to SS and QZ, ZIA HD009001-01 to ADD). This research was supported by the Intramural Research Program of the National Institutes of Health (NIH). The contributions of the NIH author(s) were made as part of their official duties as NIH federal employees, are in compliance with agency policy requirements, and are considered Works of the United States Government. However, the findings and conclusions presented in this paper are those of the author(s) and do not necessarily reflect the views of the NIH or the U.S. Department of Health and Human Services.

## Contributions

SS: performed the study and wrote the manuscript; ADD: designed and conducted the translational investigations, provided scientific guidance on study design, data use and result validation, and edited the manuscript; AT and AS: conducted parts of the clinical investigations, and performed data collection and curation; QZ: conceived and supervised this study and wrote the manuscript. All authors reviewed and approved the manuscript.

## Availability of data and materials

The datasets generated and analyzed during the current study are provided in the supplementary files. The R scripts used for analysis are available and publicly accessible in GitHub repository (https://github.com/ncats/drug_rep/tree/main/CLN3_Biomarker_Discovery).

## Declarations

## Ethics approval and consent to participate

Not applicable.

## Consent for publication

All authors consent to the publication of this manuscript.

