## Supplementary figures and images for "Toward Early Diagnosis and Therapeutic Discovery in CLN3 Disease: A Computational Biomarker Discovery Framework"

### Supplementary_Figure_1

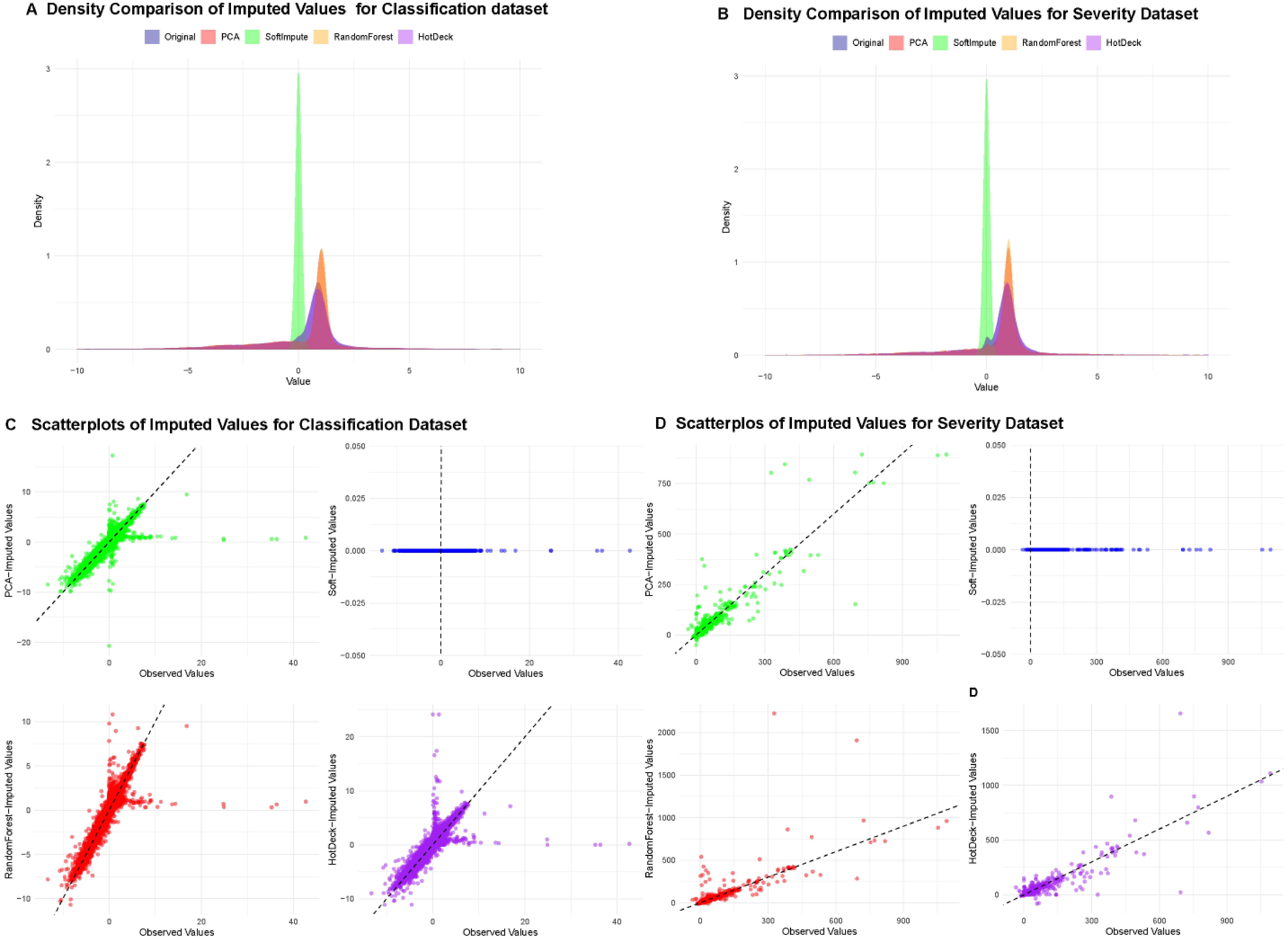

### Supplementary_Figure_2

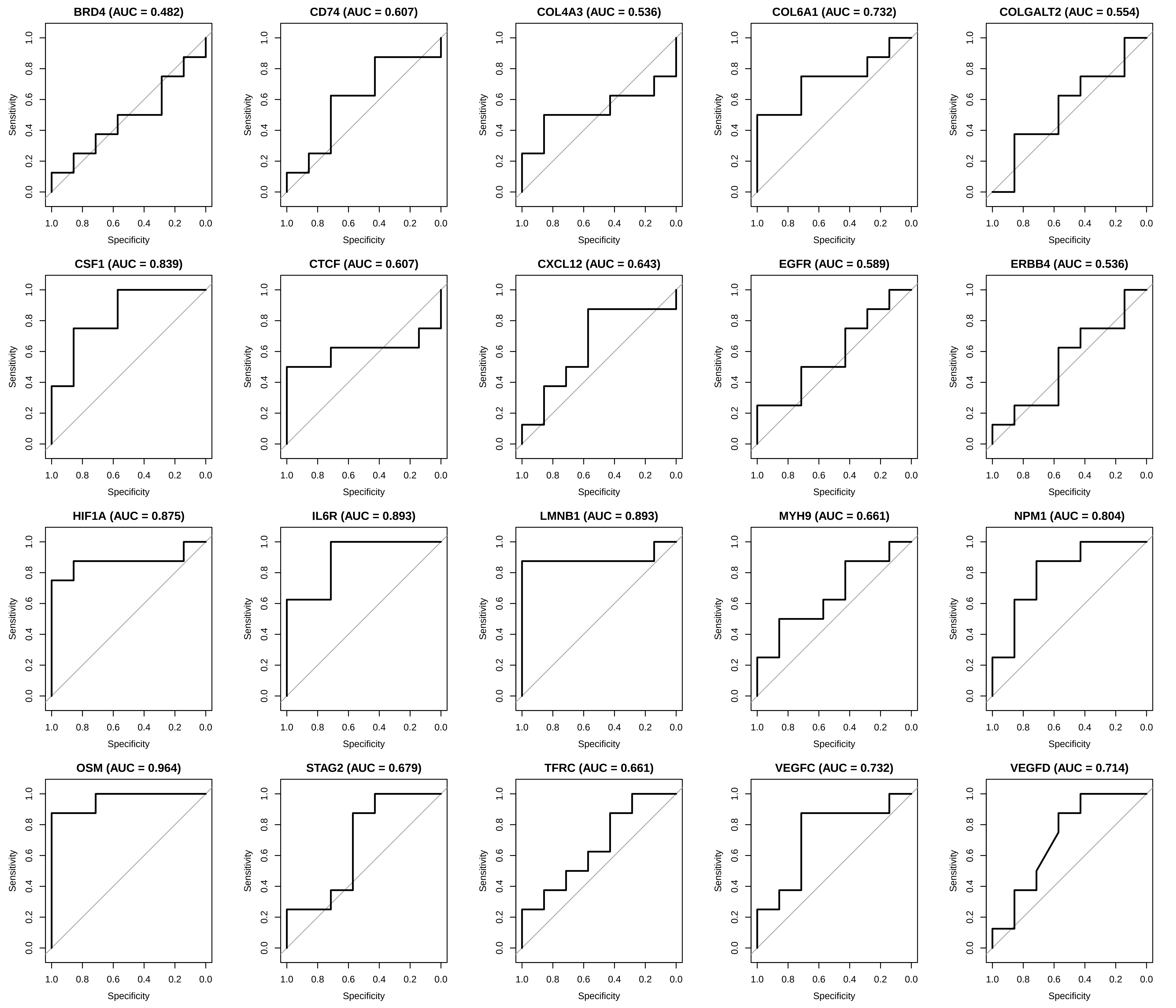
